# Approximate Reciprocal Relationship Between Two Cause-Specific Hazard Ratios in COVID-19 Data With Mutually Exclusive Events

**DOI:** 10.1101/2021.04.22.21255955

**Authors:** Sirin Cetin, Ayse Ulgen, Hakan Sivgin, Wentian Li

## Abstract

COVID-19 survival data presents a special situation where not only the time-to-event period is short, but also the two events or outcome types, death and release from hospital, are mutually exclusive, leading to two cause-specific hazard ratios (csHR_*d*_ and csHR_*r*_). The eventual mortality/release outcome can also be analyzed by logistic regression to obtain odds-ratio (OR). We have the following three empirical observations concerning csHR_*d*_, csHR_*r*_ and OR: (1) The magnitude of OR is an upper limit of the csHR_*d*_: | log(OR) | ≥ | log(csHR_*d*_)|. This relationship between OR and HR might be understood from the definition of the two quantities; (2) csHR_*d*_ and csHR_*r*_ point in opposite directions: log(csHR_*d*_)· log(csHR_*r*_) < 0; This relation is a direct consequence of the nature of the two events; and (3) there is a tendency for a reciprocal relation between csHR_*d*_ and csHR_*r*_: csHR_*d*_ ∼ 1/csHR_*r*_. Though an approximate reciprocal trend between the two hazard ratios is in indication that the same factor causing faster death also lead to slow recovery by a similar mechanism, and vice versa, a quantitative relation between csHR_*d*_ and csHR_*r*_ in this context is not obvious. These resutls may help future analyses of COVID-19 data, in particular if the deceased samples are lacking.

## Introduction

Survival analysis studies the longitudinal event data. Regression in survival analysis investigates whether a factor contributes to the hazard (rate of risks) of the event under study. The hazard ratio (HR) is the ratio of two hazards, one with the factor taking the at-risk value (e.g. smoking) and another without (e.g. not-smoking). Since hazard and HR describes the instantaneous risk, or rate of event occurrence (e.g. death from a specific disease), it is a different concept from the life-long risk of having that event (Sutradhar and Austin, 2018). As a result, regression in survival analysis (e.g. Cox regression) is different from the static case-versus-control regression analysis (e.g. logistic regression). Take the following two statements as example: “smoking makes lung cancer patients die faster” and “smoking makes a person more likely to die from lung caner than a non-smoker”; the first would be a conclusion from a survival analysis, whereas the second from a case-control type of analysis.

The COVID-19 pandemic since 2020 (Huang et al., 2020; Xu et al., 2020) provides a unique longitudinal event data. First of all, a COVID-19 patient admitted to a hospital sees his/her outcome relatively quickly: either the patient survived or not in a matter of days. As a result, there are very few right-censored data where the outcome is still unknown at the time of data collection. Of course, there exist chronic or long COVID-19 survivors who are not completely cured (Carfi et al., 2020; Rubin, 2020), but they are unlikely to die from COVID-19 in the future.

The second feature of COVID-19 longitudinal event data is that the two events, death and release from the hospital, are not the traditional “competing risk events” (Austin et al., 2016; Austin and Fine, 2017). Although not strictly defined as such, competing risks events are often two unfavorable events with one occurring before the occurrence of another. In COVID-19 data, the event of being released from hospital is a favorite event, and a description of them in principle should not be connected to words like “risk” or “hazard”. A higher HR for the event of releasing from hospital implies a faster recover, thus a factor that contributes to this higher HR actually provide protection.

Also, in the COVID-19 data, the event of death and the event of released from hospital are mutually exclusive. Although organ transplant and death can be mutually exclusive when the organ involved is (e.g.) heart, transplant of many other types of organ is an event that preceed, and may have an impact on, death. Even in the case of heart transplant, survival after the operation is not completely guaranteed. Regardless of these detail, factors affecting transplant timing are basically external, whereas those affecting mortality without a transplant are mostly internal. Our COVID-19 death/release mutually exclusive event pair does not have a correspondence in death/organ-transplant pairs.

As discussed in (Pintilie, 2007; Austin et al., 2016), given the the event time (*T, k*) where *T* is the time to event (or to censoring), *k*=1,2 for two event types (we may understand that *k* = 0 means right-censored), there are two ways to define hazard function: (1) cause-specific hazard function,

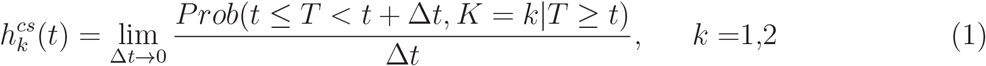

and (2) subdistribution hazard function (or Fine-Grey model),

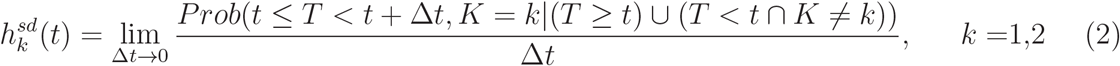

where *K* for event type (1 or 2), and conditioning *T* ≥ *t* means the function is defined before the occurring of the event, and the added conditioning (*T < t*) ∩ (*K* = *k*) means event of alternative type may be allowed to occur before. In the first approach (cause-specific), all alternative events are converted to right-censored (i.e., *k* = 0).

Because the event of dead and release are mutually exclusive, we cannot use subdistribution hazard function. We can indeed study the two cause-specific hazard functions or HRs, one for time-to-death (treating release event as right-censored) and another for time-to-release (then treating death event as right-censored):

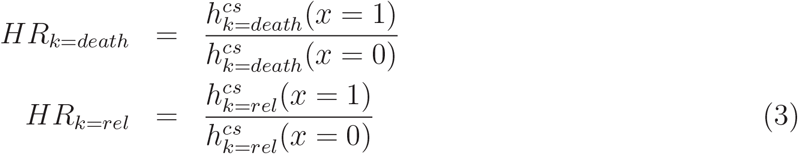

where *x* is the independent binary factor (for continuous factor, the definition is similar, with two hazards evaluated at *x*-levels differing by 1), and the time dependency is supposedly canceled. The question we ask: what is the relationship between the two cause-specific hazard ratios, csHR_*k*=*death*_ and csHR_*k*=*rel*_? Intuitively, if a factor value leads to faster death, the same factor may lead to slow recovery, and vice versa. In other words, a larger csHR_*k*=*death*_ may imply a smaller csHR_*k*=*rel*_. Our working hypothesis is that csHR_*k*=*death*_= 1/csHR_*k*=*rel*_, which is also hypothesized to be equal to the odds-ratio from a logistic regression analysis. We will use a survival data of *n* = 450 COVID-19 test our working hypothesis. Previously we asked the question on whether the two survival analyses and one logistic regression can all identify a risk factor (Cetin et al., 2021). Here we are asking more quantitative questions concerning the three analyses.

## Data

The COVID-19 patients dataset (n=450) used in this study was collected from the Tokat State Hospital. Electronic medical records, including patient demographics, clinical manifestation, comorbidities, laboratory tests results were collected. According to the severity and outcomes of the patients, they were divided into two groups: deceased/nonsurviving, released/survived severe groups. Clinical and laboratory data were comparable among the two groups. Ministry of Health permission and the ethics committee of Tokat GaziOsmanPasa University Ethics committee permission was obtained with the number 83116987/360 on March 4, 2021.

Besides age and gender, we selected these 18 laboratory testing measurements at the time of hospital admission in the survival analysis n value is the number of samples with measurement value): (from the complete blood count panel) white blood cell (WBC) count (n=432), neutrophil (NEU) count (n=383), lymphocyte (LYM) count (n=382), hemoglobin (HGB) (n=432), platelet (PLT) (n=432), mean corpuscle volume (MCV) (n=432), mean platelet volume (MPV) (n=430); (from metabolic panel) glucose (n=449), alanine adinotransferase (ALT) (n=435), aspartate aminotrasferase (AST) (n=429), blood urea nitrogen (urea or BUN) (n=436), creatine (n=436), calcium (n=393), potassium (n=433), sodium (n=435); (others) ferritin-1 (FER1) (n=407), d-dimer (n=390), and lactate dehydrogenase (LDH) (n=316). The NLR (neutrophil/lymphocyte ratio) and PLR (platelet/lymphocyte ratio) are derived quantities. There are other test measurements, either only available for fewer number of patients, or due to other reasons, that are not included in the analysis.

The following variables seem to follow a normal distribution after the log transformation better than the original values: ferritin-1, d-dimer, glucose, ALT, AST, urea, creatine, LDH, WBC, NEU, and perhaps LYM and PLT.

## Method

The alternative use of Cox regression for cause-specific hazard is applied:

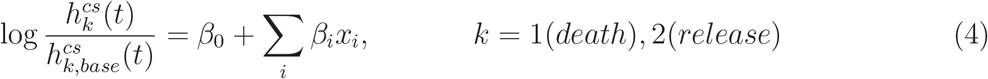

where 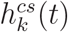 is the cause-specific hazard function defined in Eq.1, 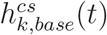 is an undefined baseline hazard function. The left-hand side of Eq.4 contains time *t* whereas the right-hand side does not, which is the proportional hazard assumption, that the time-dependent part is cancelled. {*x*_*i*_} is a list of factors. The single-variate Cox regression limits index *i* to only one variable. In practice, cause-specific Cox regression is carried out by labeling event=2 as right-censored event=0 when focusing on event=1, and vice versa when focusing on event=2.

## Results

### Overview of the survival analysis and logistic regression analysis results

Large number of analysis run results are included in Tables 1,2,3. Table 2 contains factors in a metabolic panel, Table 3 are factors in a complete blood count panel, and Table 1 lists the rest. The first column is the result from single-factor Cox regression survival analysis using death as event of interest and release as right-censored event. The second column is the Cox regression results by switching the two events. The listed results include the cause-specific hazard ratio (csHR) and its 95% confident interval (CI), and the *p*-value for testing csHR equal to 1. The third column is the logistic regression result comparing the dead and release samples, where the results include odds-ratio (OR) and its 95% CI, and *p*-value for testing OR=1.

**Table 1:** Results from two cause-specific survival analyses and logistic regression analysis with a single factor: gender, age, FER1, d-dimer, and LDH. The cs*HR*_*d*_ (95%CI) is the cause-specific hazard-ratio for time-to-death event and its 95% confidence interval; *pv*_*d*_ is the corresponding *p*-value; cs*HR*_*r*_ and *pv*_*r*_ are hazard ratio and *p*-value for time-to-release event; OR(95% CI) and pv(LR) are odds-ratio (and its 95% confidence interval) and *p*-value from logistic regression. *P*-values smaller than 0.005 are shown in boldface. All similar results for log-transformed factor value and discretized (binarized) factors are also shown, where the threshold used to discretization are given in the first column.

| factor | csHR<sub>d</sub> (95% CI) | pv<sub>d</sub> | csHR<sub>r</sub> (95% CI) | pv<sub>r</sub> | OR (95% CI) | pv(LR) |
| --- | --- | --- | --- | --- | --- | --- |
| age | 1.052 (1.033, 1.072) | **9e-8** | 0.973 (0.968, 0.978 ) | **3.5e-26** | 1.076 (1.054, 1.099) | **9.4e-12** |
| ≥70 | 3.705 (2.259, 6.078) | **2.1e-7** | 0.461 (0.368, 0.577) | **1.4e-11** | 7.69 (4.51, 13.09) | **6e-14** |
| gender | 1.048 (0.689, 1.594) | 0.83 | 1.381 (1.118, 1.706) | **0.0028** | 0.754 (0.475, 1.197) | 0.23 |
| FER1 | 1.0007 (1.0004, 1.001) | **3.1e-7** | 0.9987 (0.9984, 0.999) | **3.1e-18** | 1.002 (1.001, 1.002) | **4.3e-19** |
| log(FER1) | 1.703 (1.395, 2.077) | **1.6e-7** | 0.645 (0.6, 0.693) | **2.6e-33** | 3.108 (2.402, 4.022) | **6.5e-18** |
| >782.9 | 2.66 (1.751, 4.039) | **4.5e-6** | 0.26 (0.182, 0.371) | **1.1e-13** | 10.88 (6.35, 18.64) | **3.5e-18** |
| d-dimer | 1.558 (1.356, 1.789) | **3.6e-10** | 0.561 (0.486, 0.647) | **2.1e-15** | 2.501 (2.051, 3.049) | **1.3e-19** |
| log(dd) | 1.938 (1.552, 2.419) | **5.2e-9** | 0.571 (0.517, 0.632) | **1e-27** | 3.347 (2.545, 4.402) | **5.4e-18** |
| > 1.36 | 3.259 (2.11, 5.034) | **1e-7** | 0.319 (0.232, 0.439) | **2.2e-12** | 10.17 (6.007, 17.220) | **6e-18** |
| LDH | 1.0001 (1, 1.0002) | 0.03 | 0.997 (0.996, 0.998) | **8.6e-10** | 1.008 (1.006, 1.01) | **4.4e-13** |
| log(LDH) | 1.884 (1.574, 2.254) | **4.7e-12** | 0.298 (0.214, 0.415) | **7.1e-13** | 35.22 (14.89, 83.30) | **5.1e-16** |
| > 406.5 | 4.975 ( 3.152, 7.852 ) | **5.5e-12** | 0.235 ( 0.154, 0.36 ) | **2.3e-11** | 21.15 (11.37, 39.35) | **5.6e-22** |

**Table 2:**
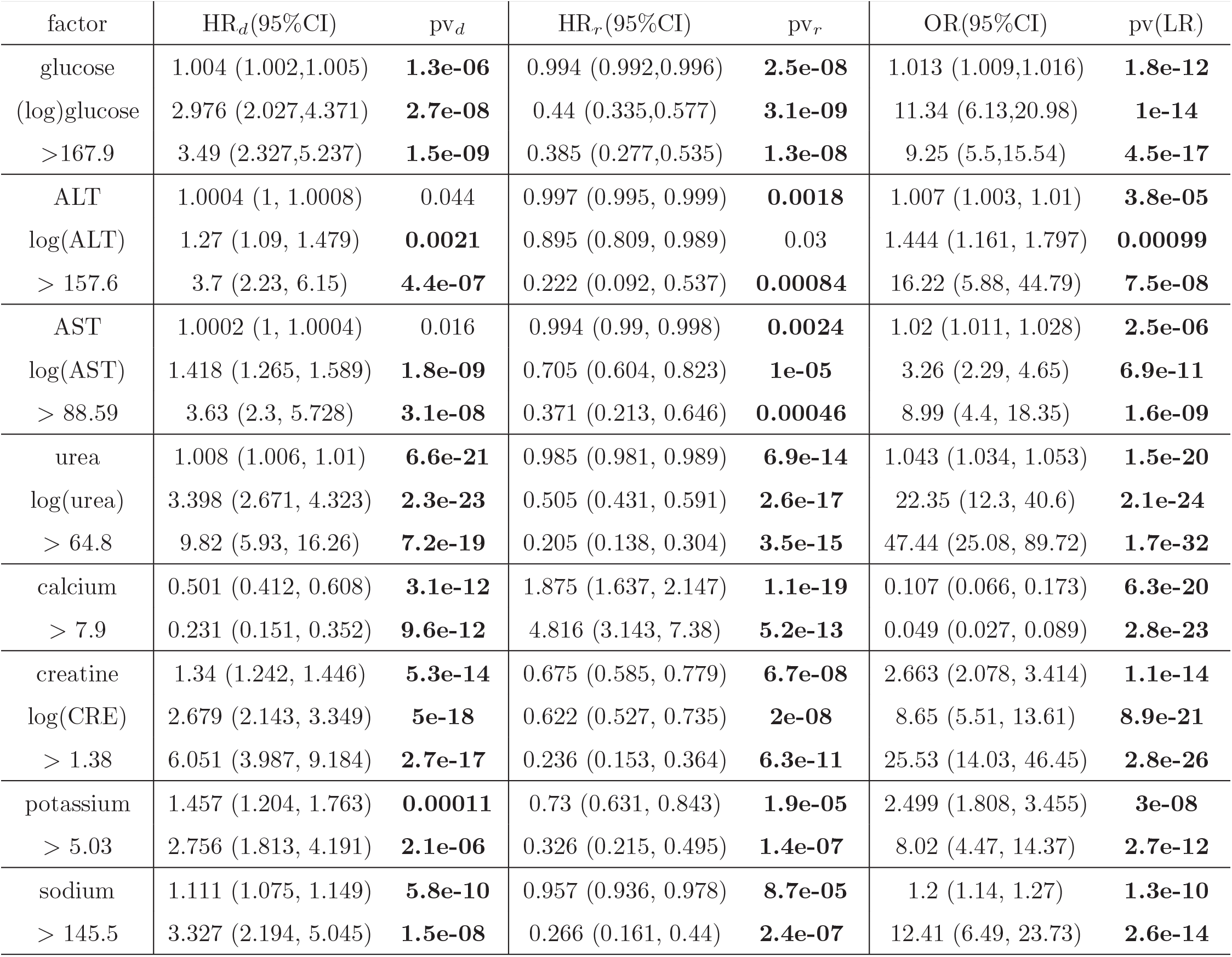
Similar to Table 1 but for factors measured by a metabolic panel blood test.

| factor | HR<sub>d</sub>(95%CI) | pv<sub>d</sub> | HR<sub>r</sub>(95%CI) | pv<sub>r</sub> | OR(95%CI) | pv(LR) |
| --- | --- | --- | --- | --- | --- | --- |
| glucose | 1.004 (1.002,1.005) | **1.3e-06** | 0.994 (0.992,0.996) | **2.5e-08** | 1.013 (1.009,1.016) | **1.8e-12** |
| (log)glucose | 2.976 (2.027,4.371) | **2.7e-08** | 0.44 (0.335,0.577) | **3.1e-09** | 11.34 (6.13,20.98) | **1e-14** |
| >167.9 | 3.49 (2.327,5.237) | **1.5e-09** | 0.385 (0.277,0.535) | **1.3e-08** | 9.25 (5.5,15.54) | **4.5e-17** |
| ALT | 1.0004 (1, 1.0008) | 0.044 | 0.997 (0.995, 0.999) | **0.0018** | 1.007 (1.003, 1.01) | **3.8e-05** |
| log(ALT) | 1.27 (1.09, 1.479) | **0.0021** | 0.895 (0.809, 0.989) | 0.03 | 1.444 (1.161, 1.797) | **0.00099** |
| > 157.6 | 3.7 (2.23, 6.15) | **4.4e-07** | 0.222 (0.092, 0.537) | **0.00084** | 16.22 (5.88, 44.79) | **7.5e-08** |
| AST | 1.0002 (1, 1.0004) | 0.016 | 0.994 (0.99, 0.998) | **0.0024** | 1.02 (1.011, 1.028) | **2.5e-06** |
| log(AST) | 1.418 (1.265, 1.589) | **1.8e-09** | 0.705 (0.604, 0.823) | **1e-05** | 3.26 (2.29, 4.65) | **6.9e-11** |
| > 88.59 | 3.63 (2.3, 5.728) | **3.1e-08** | 0.371 (0.213, 0.646) | **0.00046** | 8.99 (4.4, 18.35) | **1.6e-09** |
| urea | 1.008 (1.006, 1.01) | **6.6e-21** | 0.985 (0.981, 0.989) | **6.9e-14** | 1.043 (1.034, 1.053) | **1.5e-20** |
| log(urea) | 3.398 (2.671, 4.323) | **2.3e-23** | 0.505 (0.431, 0.591) | **2.6e-17** | 22.35 (12.3, 40.6) | **2.1e-24** |
| > 64.8 | 9.82 (5.93, 16.26) | **7.2e-19** | 0.205 (0.138, 0.304) | **3.5e-15** | 47.44 (25.08, 89.72) | **1.7e-32** |
| calcium | 0.501 (0.412, 0.608) | **3.1e-12** | 1.875 (1.637, 2.147) | **1.1e-19** | 0.107 (0.066, 0.173) | **6.3e-20** |
| > 7.9 | 0.231 (0.151, 0.352) | **9.6e-12** | 4.816 (3.143, 7.38) | **5.2e-13** | 0.049 (0.027, 0.089) | **2.8e-23** |
| creatine | 1.34 (1.242, 1.446) | **5.3e-14** | 0.675 (0.585, 0.779) | **6.7e-08** | 2.663 (2.078, 3.414) | **1.1e-14** |
| log(CRE) | 2.679 (2.143, 3.349) | **5e-18** | 0.622 (0.527, 0.735) | **2e-08** | 8.65 (5.51, 13.61) | **8.9e-21** |
| > 1.38 | 6.051 (3.987, 9.184) | **2.7e-17** | 0.236 (0.153, 0.364) | **6.3e-11** | 25.53 (14.03, 46.45) | **2.8e-26** |
| potassium | 1.457 (1.204, 1.763) | **0.00011** | 0.73 (0.631, 0.843) | **1.9e-05** | 2.499 (1.808, 3.455) | **3e-08** |
| > 5.03 | 2.756 (1.813, 4.191) | **2.1e-06** | 0.326 (0.215, 0.495) | **1.4e-07** | 8.02 (4.47, 14.37) | **2.7e-12** |
| sodium | 1.111 (1.075, 1.149) | **5.8e-10** | 0.957 (0.936, 0.978) | **8.7e-05** | 1.2 (1.14, 1.27) | **1.3e-10** |
| > 145.5 | 3.327 (2.194, 5.045) | **1.5e-08** | 0.266 (0.161, 0.44) | **2.4e-07** | 12.41 (6.49, 23.73) | **2.6e-14** |

**Table 3:** Similar to Table 1 but for factors measured by a complete blood count panel test.

| factor | csHR<sub>d</sub> (95% CI) | pv<sub>d</sub> | csHR<sub>r</sub> (95% CI) | pv<sub>r</sub> | OR (95% CI) | pv(LR) |
| --- | --- | --- | --- | --- | --- | --- |
| WBC | 1.08 (1.06, 1.1) | **3.2e-15** | 0.869 (0.841, 0.897) | **4.7e-18** | 1.368 (1.279, 1.464) | **6.7e-20** |
| log(WBC) | 3.996 (2.827, 5.648) | **4.3e-15** | 0.352 (0.284, 0.437) | **3.4e-21** | 28.2 (14.24, 55.84) | **9.6e-22** |
| >11.99 | 5.056 (3.306, 7.732) | **7.7e-14** | 0.217 (0.145, 0.324) | **8.8e-14** | 22.02 (12.35, 39.25) | **1e-25** |
| NEU | 1.085 (1.064, 1.105) | **5.2e-17** | 0.85 (0.819, 0.882) | **1.3e-17** | 1.421 (1.316, 1.533) | **1.9e-19** |
| log(NEU) | 3.77 (2.775, 5.131) | **2.5e-17** | 0.402 (0.337, 0.48) | **5.9e-24** | 21.52 (11.31, 40.92) | **8.2e-21** |
| >9.07 | 6.35 (3.975, 10.15) | **1.1e-14** | 0.238 (0.162, 0.35) | **2.7e-13** | 24.21 (13.31, 44.05) | **1.7e-25** |
| LYM | 0.243 (0.157, 0.375) | **1.8e-10** | 1.18 (1.082, 1.286) | **0.00017** | 0.124 (0.0709, 0.2167) | **2.3e-13** |
| log(LYM) | 0.35 (0.276, 0.442) | **2.1e-18** | 1.73 (1.461, 2.05) | **2.1e-10** | 0.095 (0.0555, 0.1626) | **9.3e-18** |
| >0.636 | 0.232 (0.152, 0.354) | **1.2e-11** | 4.669 (2.895, 7.529) | **2.6e-10** | 0.052 (0.0275, 0.0972) | **3.8e-20** |
| HGB | 0.856 (0.781, 0.938) | **0.00085** | 1.253 (1.192, 1.318) | **1.5e-18** | 0.615 (0.542, 0.697) | **3.6e-14** |
| >9.67 | 0.605 (0.397, 0.923) | 0.02 | 3.698 (2.544, 5.374) | **7.1e-12** | 0.145 (0.084, 0.251) | **5.1e-12** |
| PLT | 0.9968 (0.995, 0.9986) | **0.00064** | 1.0004 (0.9996, 1.0012) | 0.33 | 0.994 (0.991, 0.996) | **1.9e-6** |
| log(PLT) | 0.74 (0.5908, 0.927) | 0.0088 | 1.336 (1.109, 1.609) | **0.0023** | 0.27 (0.168, 0.434) | **6.1e-8** |
| >99.58 | 0.695 (0.411, 1.175) | 0.17 | 3.341 (1.915, 5.831) | **2.2e-5** | 0.166 (0.0785, 0.35) | **2.5e-6** |
| MCV | 1.048 (1.012, 1.085) | 0.0093 | 0.96 (0.943, 0.976) | **2e-6** | 1.085 (1.044, 1.128) | **3.8e-5** |
| >92.52 | 2.179 (1.433, 3.313) | **0.00027** | 0.582 (0.426, 0.795) | **0.00066** | 3.547 (2.112, 5.955) | **1.7e-6** |
| MPV | 1.322 (1.181, 1.48) | **1.2e-6** | 0.764 (0.701, 0.832) | **8.8e-10** | 2.326 (1.876, 2.884) | **1.5e-14** |
| >11.22 | 2.45 (1.629, 3.685) | **1.7e-5** | 0.381 (0.272, 0.533) | **1.8e-8** | 6.959 (4.131, 11.722) | **3e-13** |
| NEU/LYM | 1.023 (1.0179, 1.028) | **5.7e-20** | 0.908 (0.886, 0.931) | **3.4e-14** | 1.219 (1.166, 1.274) | **1.6e-18** |
| log(NLR) | 2.721 (2.234, 3.315) | **2.5e-23** | 0.556 (0.496, 0.624) | **1.4e-23** | 10.471 (6.406, 17.115) | **7.4e-21** |
| > 9 | 10.54 (5.95, 18.69) | **7.3e-16** | 0.182 (0.121, 0.272) | **1.3e-16** | 51.64 (26.04, 102.42) | **1.5e-29** |
| PLT/LYM | 1.0003 (1.0002, 1.0004) | **1.8e-5** | 0.998 (0.997, 0.999) | **3.6e-6** | 1.004 (1.0026, 1.0054) | **1.5e-8** |
| log(PLR) | 1.871 (1.472, 2.377) | **3.1e-7** | 0.82 (0.721, 0.934) | **0.0027** | 2.41 (1.70, 3.42) | **7.9e-7** |
| >330 | 3.698 (2.417, 5.656) | **1.6e-9** | 0.43 (0.3, 0.616) | **4.1e-6** | 6.77 (3.93, 11.68) | **6.2e-12** |

We also run the same group of analysis on log-transformed factor values, if that factor better follows a normal distribution after log transformation. Then, we run the same set of analysis, when the factor value is continuous, for binarized factors with optimally chosen threshold values. Both these runs will be discussed in detail later. The reason for running large number of analyses is not for “fishing expedition” in order to have a better chance to find statistically significant results, but to test the robustness of the results. Therefore, it is not necessary to do a multiple testing correction.

We mark those *p*-values that are smaller the 0.005 in Tables 1,2,3. The reason to use 0.005 instead of 0.01 or 0.05 is explained in (Ioannidis, 2018; Colquhoun, 2017), and the practice of always adding a level when using the word “significant” is proposed in (Wasserstein et al., 2019); see also (Li et al., 2021)). Strikingly, almost all factors significantly (at 0.005 level) influence the rate of event of COVID-19 patients, and are significantly different between the deceased and survived group.

### Relationship between HR_*d*_ and HR_*r*_ for continuous factors

Another striking observation is the relationship between the two cause-specific hazard ratios. Denote csHR_*d*_ for csHR of the event of death from COVID-19 and csHR_*r*_ for csHR of the event of COVID-19 patient releasing from hospital. it can be easily seen from Table 1,2,3 that if csHR_*d*_ *>* 1, the corresponding csHR_*r*_ *<* 1, and vice versa. A simple mathematical expression of this fact is:

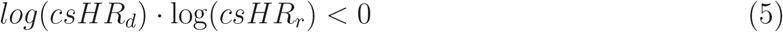

The only exception is the factor of gender. But the 95%CI is so large to have both *<* 1 and *>* 1 values, it should not really be considered as an exception. The opposite direction of csHR_*d*_ and csHR_*r*_ is understandable: a risk factor for faster death in a deceased patient would also make a surviving patient recover longer.

Table 1,2,3 seem to also show that the larger csHR_*d*_, and smaller csHR_*r*_, and vice versa. In order to check if there is a numerical relationship between csHR_*d*_ and csHR_*r*_, we plot 1/HR_*r*_ as function of csHR_*d*_ in Fig.1. The line csHR_*d*_ × csHR_*r*_ = 1 is marked by the slope=1 line in Fig.1. There are many factors clustered near the csHR_*d*_=csHR_*r*_=1 point and a close-up plot is shown separately. A factor is labeled in red if *p*-values for both csHR is significantly (at level 0.005) different from 1, and in blue if one or both csHR is not significant. Our working hypothesis can be written as a reciprocal relation between csHR_*d*_ and csHR_*r*_:

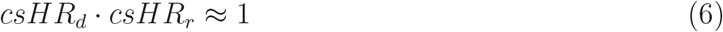

**Figure 1:**
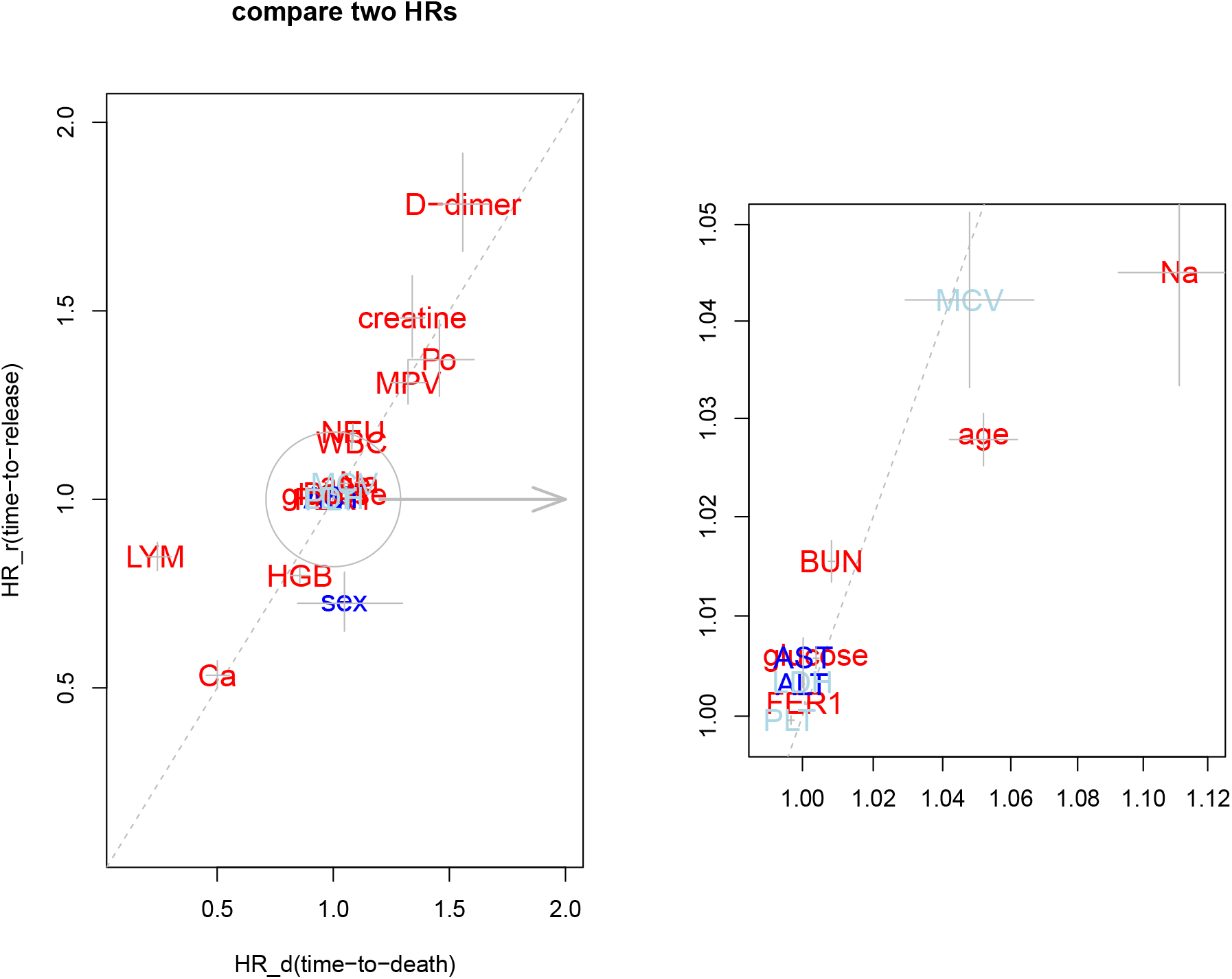
The x-axis is the cause-specific hazard ratio csHR_*d*_ for death event, and y-axis is reciprocal of the hazard ratio for the release event (1/HR_*r*_), for the 18 blood test measurements as well as age and gender. The diagonal line indicates the exact relationship csHR_*d*_=1/csHR_*r*_. A factor is in red if its *p*-values (for testing csHR_*d*_ and csHR_*r*_=1) are both smaller than 0.005; in blue if both *p*-values are larger than 0.005; and light-blue if one of the two *p*-values is smaller than 0.005. Because there are many factors having HR close to 1, the right subplot presents a close-up near csHR_*d*_=csHR_*r*_=1.

### Relationship between hazard (rate) ratio and odds (risk) ratio

As emphasized in (Sutradhar and Austin, 2018), survival analysis estimates the relative rates (of risk) whereas case-control type of analysis such as logistic regression estimates the relative (static or cumulative) risks. Table 1,2,3 show that OR seems to have a larger magnitude than csHR_*d*_. Fig.2 shows y=OR as a function of x= csHR_*d*_. Unlike Fig.1 where dots are scattered near the slope=1 line, in Fig.2, dots systematically deviates from the diagonal line. In fact, if a factor is a risk (OR *>* 1 and csHR_*d*_ *>* 1), we have OR *>* csHR_*d*_, and if a factor is a protection (OR *<* 1 and csHR_*d*_ *<* 1), then OR *<* csHR_*d*_. We can summarize these into a working hypothesis:

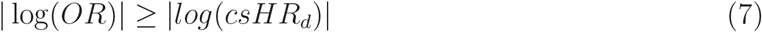

**Figure 2:**
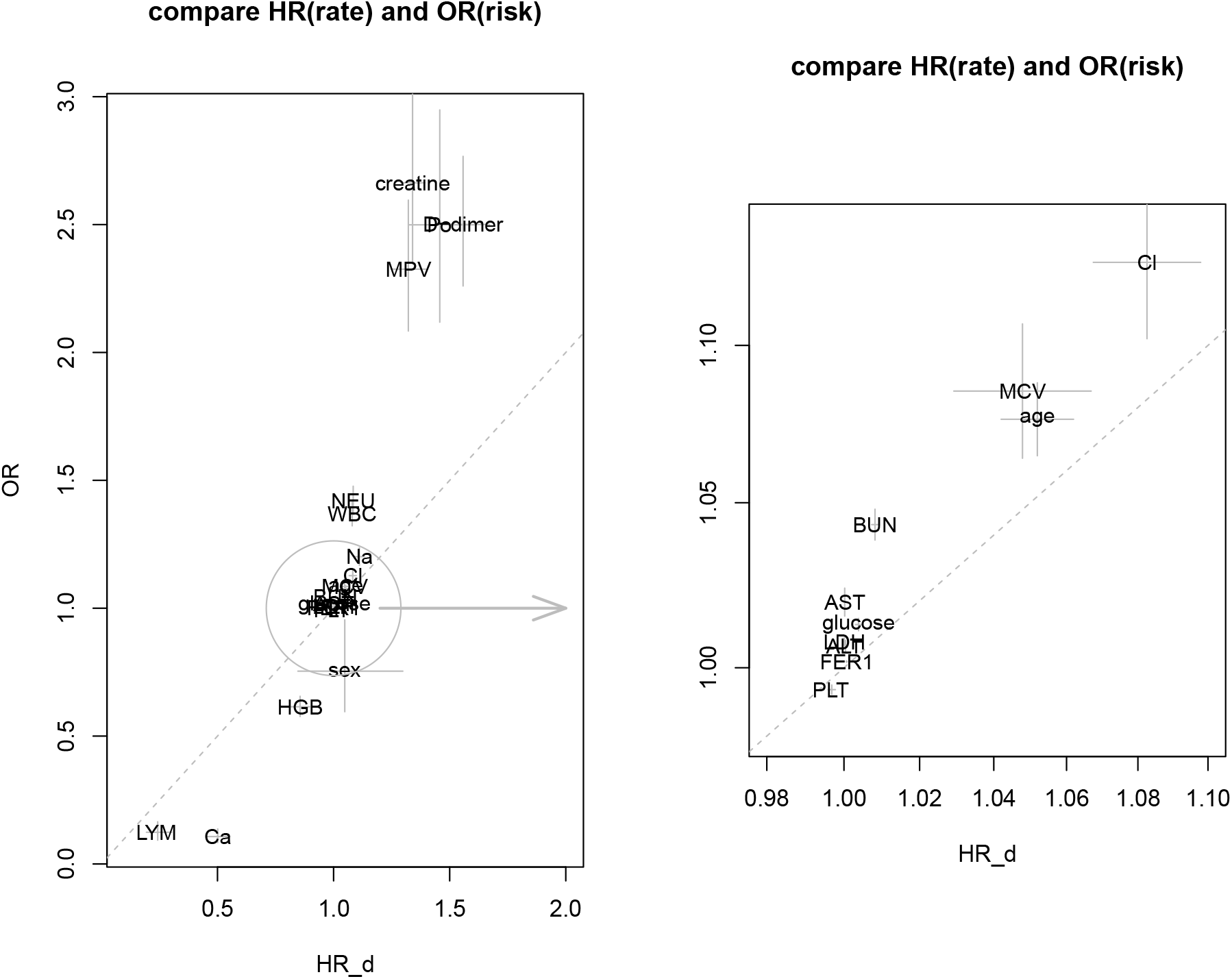
The x-axis is the cause-specific hazard ratio csHR_*d*_ for death event, and the y-axis the odds-ratio (OR) from logistic regression, for 20 factors. The right subplot presents a close-up near csHR_*d*_=OR=1.

Eq.7 might be proved in a simple approximation as follows: the risk function *F* (*t*) is known to be related to hazard rate function *h*(*t*):

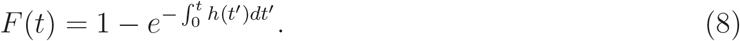

Therefore, the odds-ratio is (the subscript 1,2 refers to two states in a binary variable or a two numerical level of a continuous factor with one unit difference, and *not* refers to the two competing-risks):

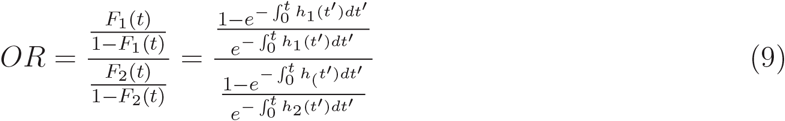

In the proportional hazard assumption, *h*_1_(*t*) = *α*_1_*h*_0_(*t*), *h*_2_(*t*) = *α*_2_*h*_0_(*t*), where *h*_0_(*t*) is a baseline hazard function, and denote 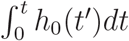 as *H* (*t*), Eq.9 becomes

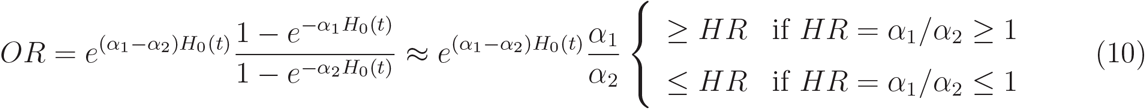

One approach in getting the approximation in Eq.10 is the Taylor expansion assuming small *H*_0_(*t*) (Stare, 2016).

### Relationship between csHR_*d*_ and csHR_*r*_ for binarized continuous factors

The HR for continuous factors measures the ratio of two hazard rates when the unit of the factor increases by one. Therefore, when one unit change is negligible compared to the possible range, HR can be very close to 1. In order to see the true impact of a continuous variable, we discretize continuous factors into levels. Although one can choose three levels for below-normal, normal, and above-normal, the normal range of a factor may not be universally accepted.

We use binary levels (higher and lower than a threshold) where the threshold value is a compromise between two selections: the first threshold is chosen to maximize the Youden index (Youden, 1950), which is simply the sum of sensitivity and specificity (minus 1). The second threshold is chosen by providing the population prevalence of cases (which is set at 10%), which in turn gives weight to samples in the dataset. Both thresholds are obtained from the *medcalc*.*org* program. The final threshold is a geometric mean of the Youden-based threshold and that after considering the 10% population case prevalence. The resulting threshold values for all factor (except for age, neutrophil/lymphocyte ratio, platelet/lymphocyte ratio, where the thresholds are more intuitively selected) are given in Tables 1,2, 3. The resulting csHR_*d*_ and csHR_*r*_ have larger magnitude than the corresponding continuous value, because the “unit change” is much larger.

In order to study the numerical relationship between csHR_*d*_ and csHR_*r*_ for binarized factors, we examine all possible threshold values and plot csHR_*d*_ (red) and 1/csHR_*d*_ (blue) as a function of threshold value, for 18 test measurements, in Fig.3. The 95% confidence interval (CI) of csHR_*d*_ or 1/csHR_*r*_ is marked with dash vertical lines. If the discretized factor is not significant at a corresponding threshold, red dots turn pink and blue dots turn light-blue. We also mark normal ranges of blood tests from two different sources (as grey horizontal lines) and the threshold used in Table 1 (as downward arrow in grey). We consistently found the 95% CI of csHR_*d*_ and 1/csHR_*r*_ overlap with each other at the chosen (optimal) threshold value. In other words, when a reasonable threshold value is used to convert a continuous factor to a binary factor, and running two survival analyses results in a roughly reciprocal relationship between the two cause-specific hazard ratios.

**Figure 3:**
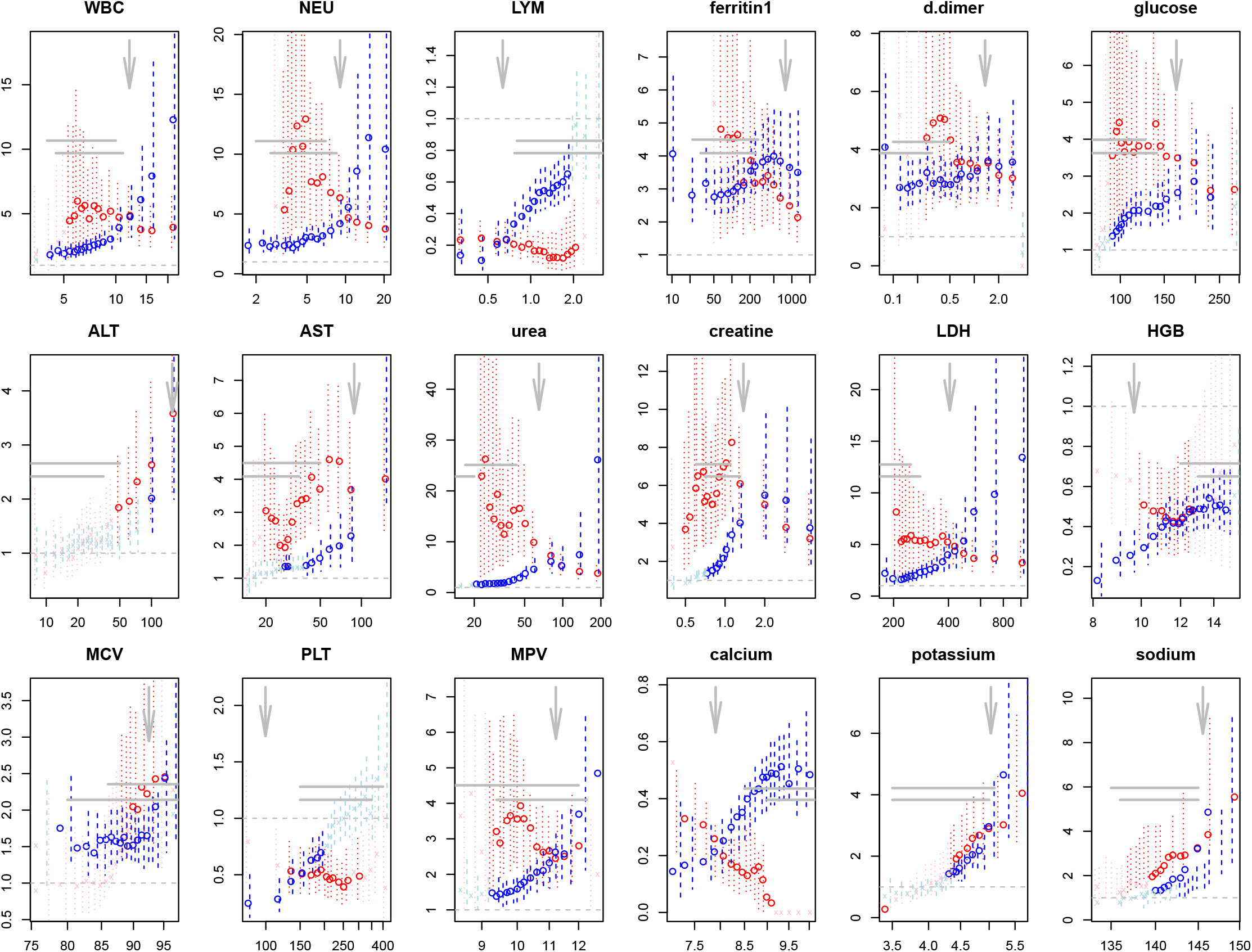
The csHR_*d*_ (red) and 1/csHR_*r*_ (blue) for binarized 18 blood test measurements as a function of the threshold used to discretize these factors. The 95% CI of csHR_*d*_ or 1/csHR_*r*_ are shown in dashed vertical lines. If the discretized factor’s csHR is not significant (at 0.01 level) by the survival analysis, its color turns from red (blue) to pink(light-blue). The threshold used in Table 1 is shown as a downward arrow. The two horizontal lines represent the normal range of these blood test results (from two different sources), and horizontal dash line is csHR=1.

### Relationship between csHR_*d*_ and OR for binarized continuous factors

Similar to Fig.2 where we show the scatter plot for cause-specific hazard ratio for time-to-death (x-axis) and logistic regression odds-ratio (y-axis), Fig.4 shows the similar scatter plot for the discretized factors. The 95% CI for both are shown by horizontal and vertical segments. All points in the first quadrant are above the diagonal line, and those in the third quadrant below the diagonal line. Therefore, | log(*OR*)| ≥ | log(*csHR*_*d*_)| is true for all binarized factors.

**Figure 4:**
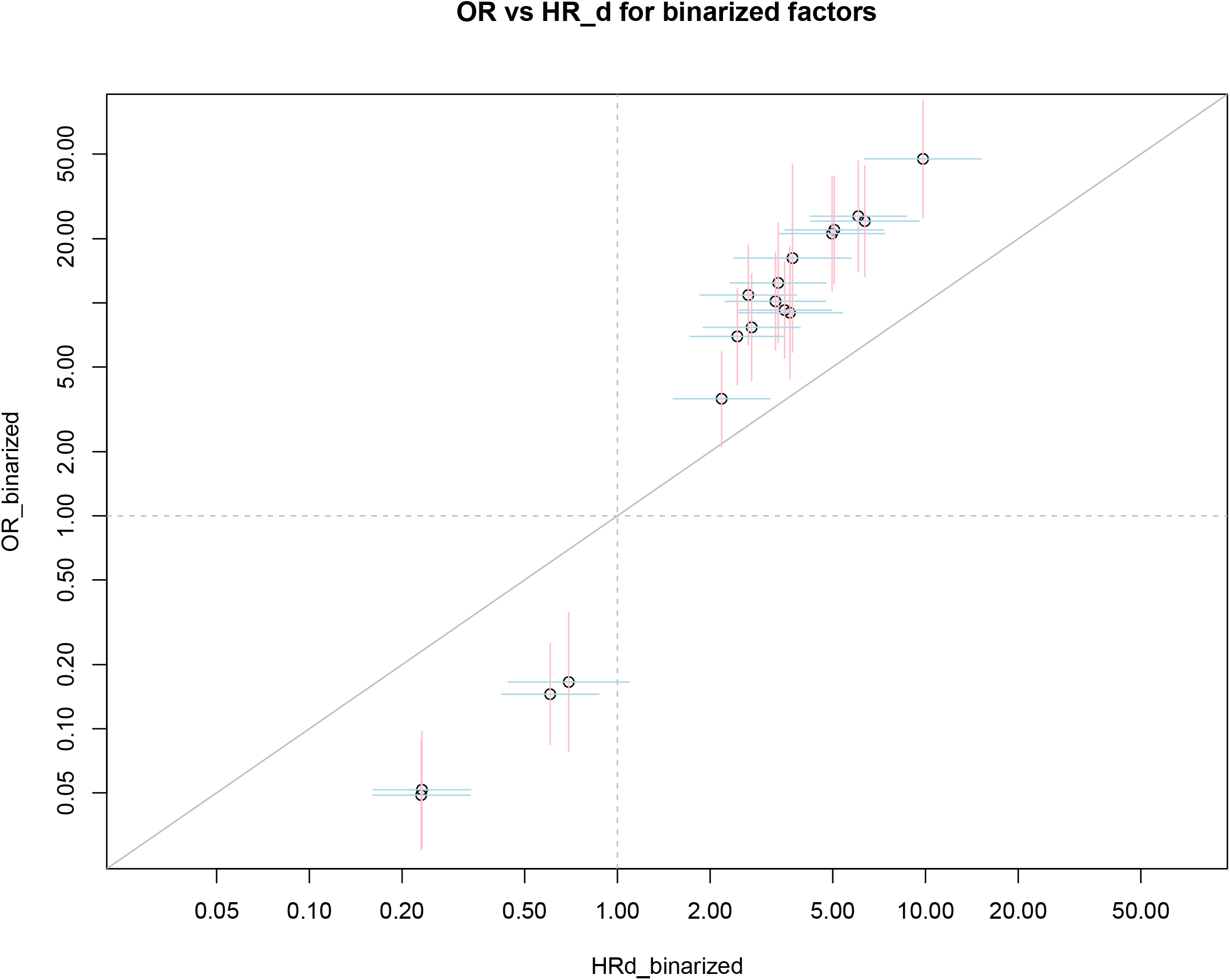
Similar to Fig.2, but for discretized/binarized factors: the x-axis is the cause-specific hazard ratio csHR_*d*_ for death event, and the y-axis the odds-ratio (OR) from logistic regression.

### Effect of log-transformation of factor values

Remember the meaning of HR for continuous variable is the ratio of two hazards evaluated at two factor values differing by one unit: *x*_1_ = *c, x*_2_ = *c* + 1. The dependence on *c* is supposed to be averaged out. If the factor is log-transformed, the two evaluation points are log(*x*_1_) = *c*′, log(*x*_2_) = *c*′ + 1, or *x*_2_ is *x*_1_ multipled by a constant 2.718. Not only this one-unit change is much larger, but also, the change in the original scale *x*_2_ − *x*_1_ = *e*^*c*′+1^ − *e*^*c*′^ = 1.718 · *e*^*c*′^ depends on *c*′. Table 1,2,3 show that csHR_*d*_ or cs*HR*_*r*_ are dramatically larger (in magnitude) than the un-logged factors. The effect of log-transformation on *p*-value is unclear, though in our examples, the test becomes more significant after the factor being log-transformed.

The concept of harzard ratio has been cautioned in (Hernán, 2010). In particular, the instantaneous incident rate for an event to occur may depend on the time, and HR is only an average over the potential time dependence. Our attempt to binarize a continuous factor or log-transform a factor illustrates a similar problem. If HR not only depends on the one-unit-change step, but also depends on which level this step is made, and depends on whether the step is in additive scale or multiplicative scale, then the average may not capture the whole spectrum of the behavior of hazard in its full range.

## Discussion

The COVID-19 time-to-event data is unique not only the two events, mortality and discharge, are mutually exclusive, but also a susceptible factor might be behind faster death and slower release at the same time. If we consider heart transplant and other open-heart operations as events that may have saved a patient’s life, thus are mutually exclusive with mortality, we can not say that the factors causing a longer waiting time until operation are the same ones causing a faster death without these operation. For this reason, we have the basis for the reciprocal cause-specific hazard (csHR) ratios hypothesis which can only be examined in data similar to COVID-19, but not in other survival data just because two events are mutually exclusive.

The possible links between the two csHR’s have at least two consequences. The first is on hospitalization stay time. If a patient has certain condition (e.g., high glucose or hyperglycemia), the larger-than-1 csHR_*d*_ implies that the patient has a higher death-rate than those with normal glucose level; on the other hand, the smaller-than-1 csHR_*r*_ value indicates the patients should survive longer. Therefore, whether hyperglycemia increases the hospital stay time or not depends on whether the patient survives or not. The hospital stay time is of interest because of the number of hospital beds is limited, and there is a need for bed management (Roimi et al., 2021).

If a factor/condition causes the severity of a COVID-19 patient, intuitively we would conclude that patients with the condition will stay in hospital longer. In reality, if the disease is too severe, the patient will stay in hospital shorter, because the patient succumbed to death faster. Among the deceased patients, we would expect the co-existence of short and long stays, while less diverging in stay time for discharged patients. Indeed, though mean of log(stay time) between the deceased and released groups is not significantly different (t-test *p*-value= 0.15, though Wilcoxon test *p*-value is 0.00034), the variances are very different (F-test *p*-value= 1.1E-8).

The second consequence that two csHRs might be related is that if we focus on time-to-release events, we could collect much more samples simply because more patients being recovered/released than deceased. In a sense, this strategy examines which factor delays the recovering time in surviving patients. Larger sample sizes would help to detect more subtle causing factors. This strategy will become more relevant if life-saving drugs for COVID-19 are developed and nobody or almost nobody die from the disease. Even in that future event, we still have surviving patients in our possession for a survival analysis.

The fact that OR *>* csHR_*d*_ if OR *>* 1 (and OR *<* csHR_*d*_ if OR *<* 1) for both continuous factors and their discretized version, seem to be a consequence of the definition of the two quantities. Although one may use this result to obtain an upper limit of csHR_*d*_, the result in Table 1,2,3 seems to indicate that the bound is not tight. In that case, if OR ≫ csHR_*d*_, OR will not be very useful in estimating the csHR_*d*_ value.

As discussed thoroughly in the literature that we can not always assume csHR (unlike subdistribution HR) is in the same direction as OR (Lau et al., 2009; Austin et al., 2016). In other words, csHR *>* 1 does not universally imply OR *>* 1. Individual csHR also can not determine the cumulative incidence function caused by multiple risks (Latouche et al., 2013). However, our results in Tables 1,2,3 show that OR and csHR_*d*_ are always in the same direction (both larger than 1, or, both smaller than 1), indicating the difference between a theoretical possibility and the reality. Drawing cumulative incidence function is also not a goal in our analysis. Considering all these, we consider the use of csHR better fitted for COVID-19 death/release survival data, than the subdistribution HR. In fact, we doubt subdistribution HR can be applied to this situation at all, because of the exclusive nature of the two events.

In conclusion, we draw attention to the connection between the two types of mutually exclusive events, mortality and discharge, in COVID-19 survival data. We also made three observations from COVID-19 data: the opposite direction between the two csHRs: *log*(*HR*_*d*_) · log(*HR*_*r*_) *<* 0, approximately reciprocal link between them *HR*_*d*_ · *HR*_*r*_ ≈ 1, and odds-ratio as an upper limit of HR_*d*_: | log(csHR_*d*_) | ≤ | log(OR) |.

## Data Availability

available upon request

## Acknowledgement

WL acknowledges the support from Robert S Boas Center for Genomics and Human Genetics, and thanks Yaning Yang for discussions.

